# Quantifying the Spectrum of Early Motor and Language Milestones in Sex Chromosome Trisomy

**DOI:** 10.1101/2024.08.16.24312065

**Authors:** Talia Thompson, Samantha Bothwell, Jennifer Janusz, Rebecca Wilson, Susan Howell, Shanlee Davis, Karli Swenson, Sydney Martin, Karen Kowal, Chijioke Ikomi, Maria Despradel, Judith Ross, Nicole Tartaglia

## Abstract

**Background and objectives:** Sex chromosome trisomy (SCT) is a common chromosomal abnormality associated with increased risks for early developmental delays and neurodevelopmental disorders later in childhood. Our objective was to quantify the spectrum of early developmental milestones in SCT. We hypothesized later milestone achievement in SCT than the general population.

**Methods:** Data were collected as part of the eXtraordinarY Babies Study, a prospective natural history of developmental and health trajectories in a prenatally identified sample of infants with SCT. Parent reported, clinician-validated, early motor and language milestones were collected at 2, 6, 12, 18, 24, and 36-months. Age distributions of milestone achievement were compared with normative data.

**Results:** In all SCT conditions, compared with normative data, there was increased variability and a later median age of skill development across multiple gross motor and expressive language milestones. Results also show a significant amount of overlap with the general pediatric population, suggesting that for many children with prenatally identified SCT, early milestones present within, or close to, the expected timeline.

**Conclusions:** As increasing numbers of infants with prenatal SCT diagnoses present at pediatric practices, we provide an evidence-based schedule of milestone achievement in SCT as a tool for pediatricians and families. Detailed data on SCT milestones can support clinical interpretation of milestone achievement. Increased variability and later median age of milestone acquisition in SCT compared to norms support consideration of all infants with SCT as high risk.

## Background

Sex chromosome trisomy (SCT) (XXY/Klinefelter syndrome, XYY/Jacob syndrome, XXX/Trisomy X) is a common chromosomal abnormality, occurring in 1 of every 500 live births.^1^ Prior SCT research, limited by ascertainment bias and small sample sizes, has provided broad descriptions of early development, including profiles of increased risk for delays in gross motor and communication^2,3^ and high rates of early intervention.^4^ Recent advances in noninvasive prenatal screening^5^ have led to increasing rates of prenatally identified SCT and subsequently a growing population of infants with a confirmed SCT diagnosis early in life. As the literature lacks concrete information on the timing of typical milestone achievement in SCT, parents and providers lack clear guidance on what to expect during a child’s early years.

Close surveillance of key developmental milestones is a critical part of pediatric care, supporting the promotion of healthy development and the early detection of potential developmental delays.^6^ However, common surveillance methods (e.g., CDC milestones^7^ checklists) may have less utility for children with genetic conditions and those at-risk for delays such as infants born prematurely. Research has shown that the timing of milestone acquisition differs from the general population in children with Down syndrome (DS),^8^ fragile X (FXS),^9^ and preterm and very low birthrate infants.^9–11^ If this is the case for SCT as well, early developmental care should go beyond surveillance and general screening to include periodic direct developmental assessment. Further, a clear understanding of when children with SCT acquire key developmental milestones is critical for setting reasonable expectations, alerting families to potential concerns, and guiding providers in their referrals for early intervention. This is especially important with the increased frequency of prenatal SCT diagnoses, as pediatricians will be responsible for developmental care in a higher number of infants with SCT presenting to their practices. Therefore, the primary purpose of this study is to fill this gap in the SCT literature with a current, evidence-informed schedule of key early gross motor and language milestone achievement for each of the SCT conditions. These findings will support a more personalized approach to monitoring and care in SCT. Comparisons with previously published normative data to the three SCT conditions will provide critical context and a richer understanding of the SCT phenotypes, and guide recommendations for early developmental care.

## Methods

Data were collected as part of the IRB-approved eXtraordinarY Babies Natural History Study, which leverages recent advances in genetic testing with a prospective investigation of the developmental and health trajectories in a prenatally identified sample of infants with SCT (ClinicalTrials.gov NCT03396562; COMIRB 17-0118; Nemours IRB# 1151006). Inclusion criteria are prenatal identification of SCT (by cfDNA, chorionic villi sampling, and/or amniocentesis) with diagnostic confirmatory karyotype (chorionic villi sampling, amniocentesis, or postnatal), English or Spanish speaking, and child age of 6 weeks to 12 months upon enrollment. Children are excluded from participation if there is a previous diagnosis of a different genetic or metabolic disorder with neurodevelopmental or endocrine involvement, prematurity less than 34 weeks gestational age, a complex congenital malformation not previously associated with SCT, history of significant neonatal complications (i.e., intraventricular hemorrhage, meningitis, hypoxic-ischemic encephalopathy), or known central nervous system (CNS) malformation identified by neuroimaging. Study visits are conducted regularly at ages 2, 6, 12, 18, 24 months, and then yearly at two sites (Colorado and Delaware) with a combination of in-person and telehealth visits. Visits include comprehensive health and developmental history, current interventions, physical examination, and a battery of developmental assessments and parent questionnaires. Participants with gestational age <37 weeks at birth were excluded from this analysis. Tartaglia et al., (2020) provides additional details on the eXtraordinarY Babies natural history study protocol.

### Developmental Milestone Measurement

Data on the timing of milestones were collected at every study visit as part of a parent completed health and development questionnaire asking parents to report the age (in months) their child achieved key developmental milestones, including eight gross motor skills (rolling front to back, rolling back to front, sitting independently, crawling, cruising, walking, running, jumping) and four expressive communication milestones (cooing, babbling, single words, 2-word phrases).

These milestones were chosen because they can be easily observed by parents within a natural setting and delays may predict other areas of known concern in older children with SCT. During the study visit, a physician then reviewed the parent questionnaire responses by interview to confirm ages and parent understanding of the milestone. If there were discrepancies between parent reported skill and the milestone achieved (for example the parent reported the infant was “sitting independently” but physician confirmed the infant was only sitting in a propped position), the physician would adjust the data on the physician data form. The physician data form was used for data analysis.

### Normative data

Each of the twelve developmental milestones collected for the study sample were compared with existing published norms. We included normative data from studies with published values for the 25^th^, 50^th^, 75^th^, and 90^th^ percentiles for the milestones of interest from the Denver II Scales,^12^ the World Health Organization (WHO) Motor Development Study,^13^ and the Primitive Reflex Profile (PRP).^14^ As normative data were not available from a single source for all twelve milestones, we used the Denver II whenever possible (sitting, walking, running, jumping, cooing, babbling, single words, 2-word phrases). For milestones that were not included on the Denver II, we used data from the WHO (crawling and cruising) and the PRP (rolling front to back, rolling back to front). As the PRP normative dataset only provided means and standard deviations, percentiles were estimated theoretically under the assumption of a normal distribution.

### Analysis

All analyses were performed in R, version 4.4.0. Descriptive summaries by SCT are presented as median [interquartile range] and N (%). Demographic differences between SCTs were tested using Kruskal-Wallis tests for continuous variables and Fisher’s-Exact tests for categorical variables. For each milestone, achieved ages earlier than the normative 2.5^th^ percentile were removed as early outliers. Normative and SCT milestone ages are visualized from their 25^th^ – 50^th^, 50^th^ – 75^th^, and 75^th^ – 90^th^ percentiles. Differences in milestones were analyzed using simulated data based on the normative percentiles, under the assumption of a non-normal distribution, and were tested with Wilcoxon Rank-Sum tests. Differences in milestones were also analyzed between children who had a history of early intervention (EI) therapies and those who did not. Exploring whether there was an overall relationship of milestone achievement with receiving EI therapy was important to ensure therapies were not significantly impacting the distribution of milestones achievement.

## Results

Participants include 298 young children with prenatally identified SCT, including 174 with XXY, 50 with XYY, and 74 with XXX. All included children had at least one milestone age reported.

Table 1 shows sample characteristics. At the time of analysis, the median age of patients included was 4.5 years with the youngest group being XYY children, with a median age of 2.6. The majority of the cohort was white (81.9%) and non-Hispanic/Latinx (83.9%). Included children had participated in the eXtraordinarY Babies study for a median of 3 years, with XXY children having participated the longest (median: 3.5 [IQR: 2.7, 3.8] years) and XYY children having participated for the shortest period of time (median: 1.2 [IQR: 0.4, 3.1] years).

**Table 1.**
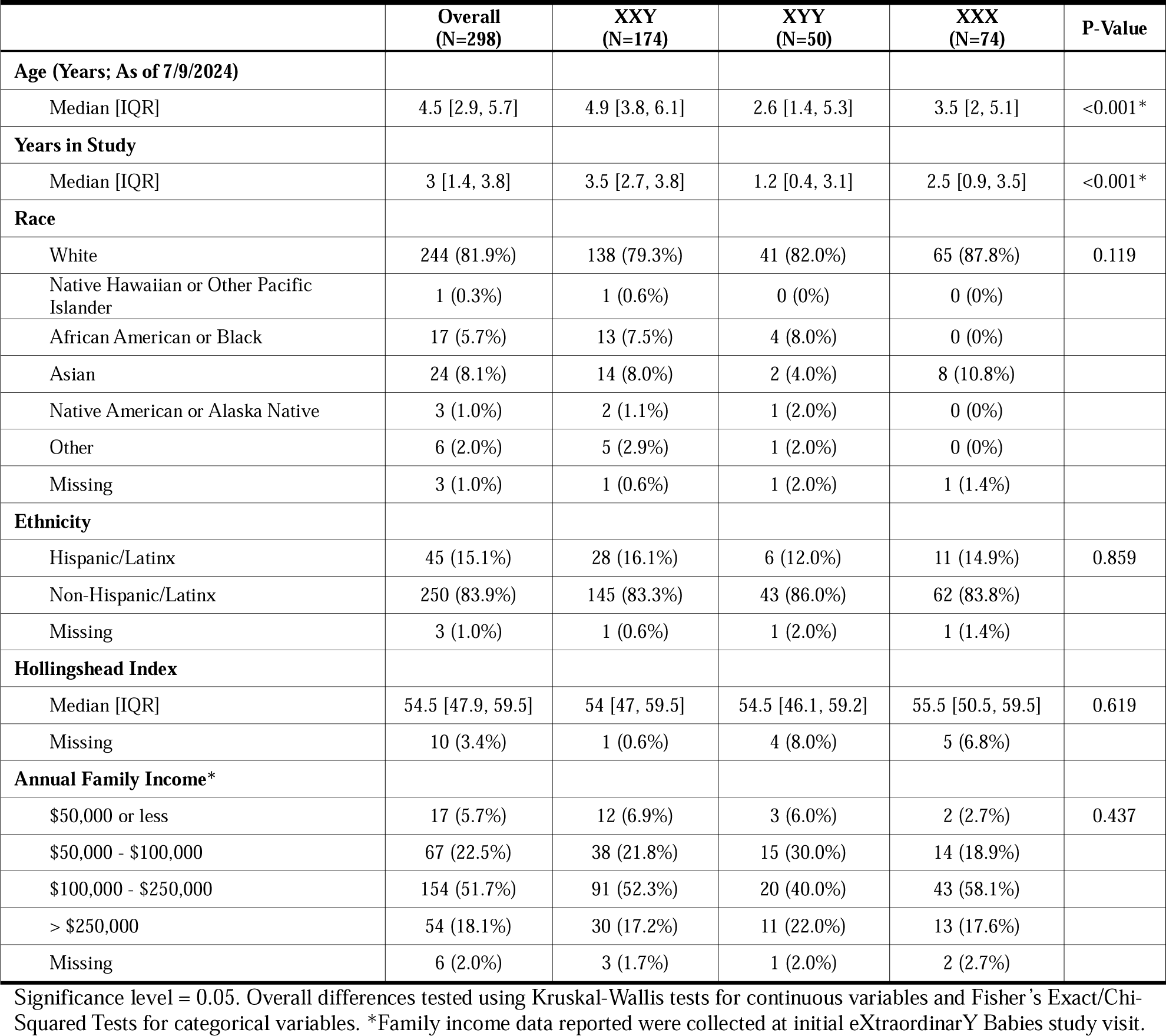
Cohort Demographics.

### Timing of Milestone Achievement in SCT Compared with Normative Datasets

Figure 1 depicts the age (in months) of milestone achievement for each SCT compared with reference norms. Age distributions are characterized by plotting the values for the 25^th^, 50^th^, 75^th^, and 90^th^ percentiles of each milestone and comparing to normative data. Results indicate that the distributions for all twelve milestones differ (later median milestone achievement; p<0.05) from the normative dataset in at least one SCT group per milestone.

**Figure 1.**
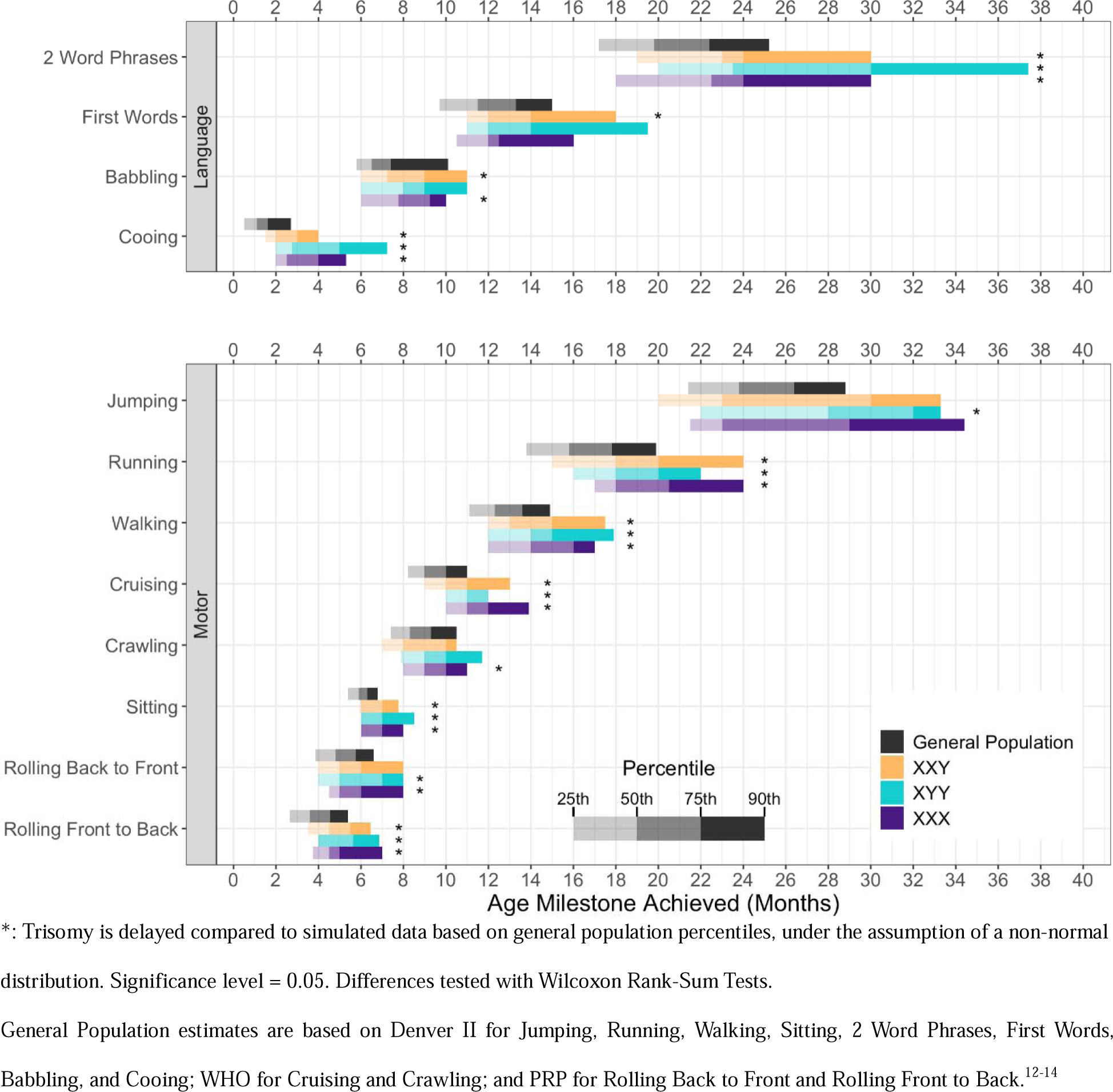
Achievement of Language and Motor milestones in Sex Chromosome Trisomy compared to the general population

### Group Differences

Table 2 shows statistical results for group differences in age of milestone achievement between the SCT conditions. Results show statistically significant group differences in cooing (p=0.005); boys with XXY achieved cooing earlier than both girls with XXX (p = 0.016) and boys with XYY (p = 0.006). Overall group differences exist for crawling (p=0.050) and cruising (p=0.012). Boys with XXY achieved crawling (p=0.017) and cruising (p=0.006) at a significantly younger age than girls with XXX. All other milestone data were statistically similar across trisomy conditions.

**Table 2.**
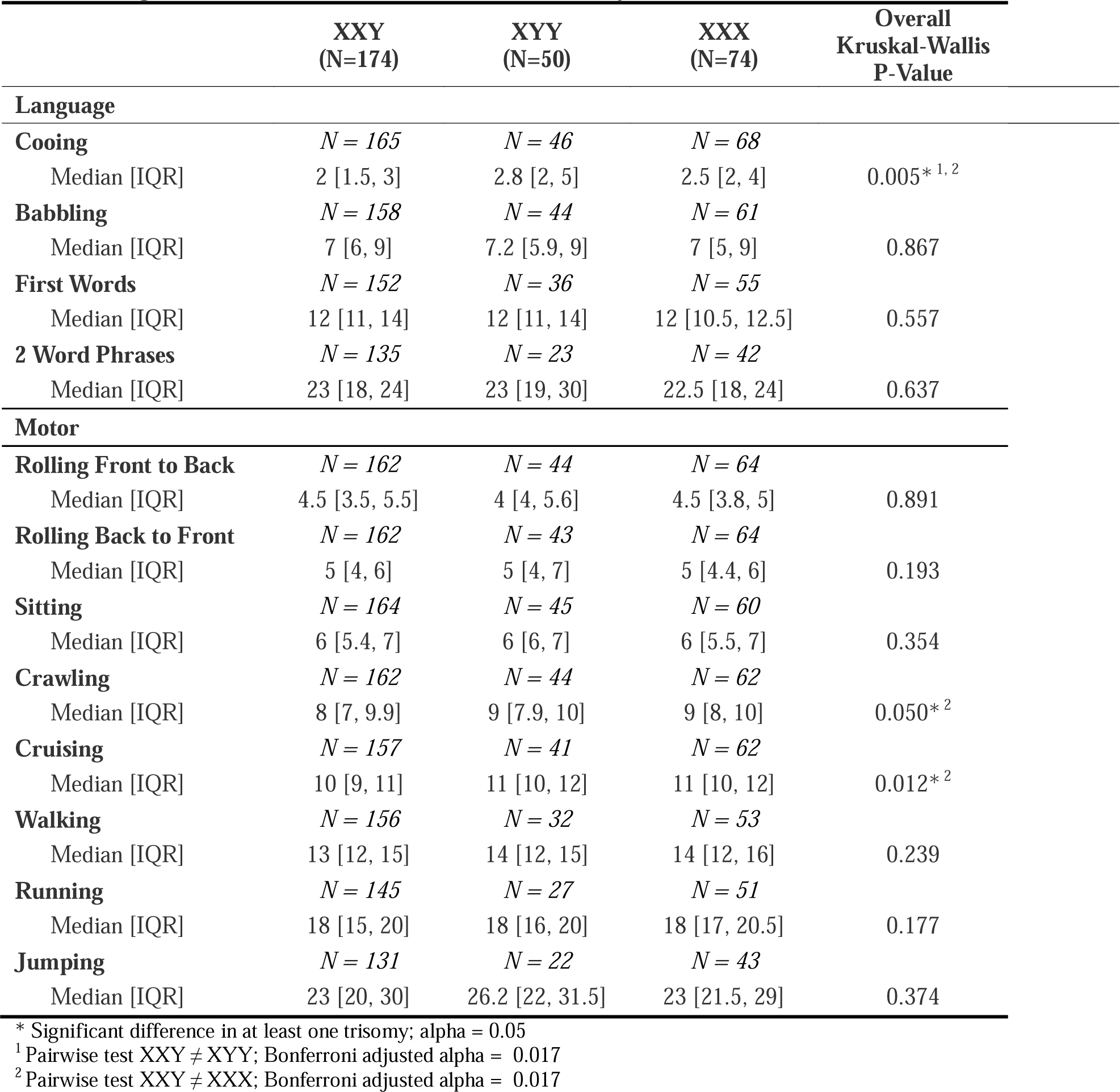
Age in Months of Milestone Achievement by SCT^1^.

### Comparisons with CDC Milestones

Table 3 shows the percent of children by SCT condition who did not achieve milestones by the age listed on the CDC milestones checklists (CDC milestones purport to represent the specific health supervision visit age when ≥75% or more of children are expected to demonstrate the skill).

**Table 3.**
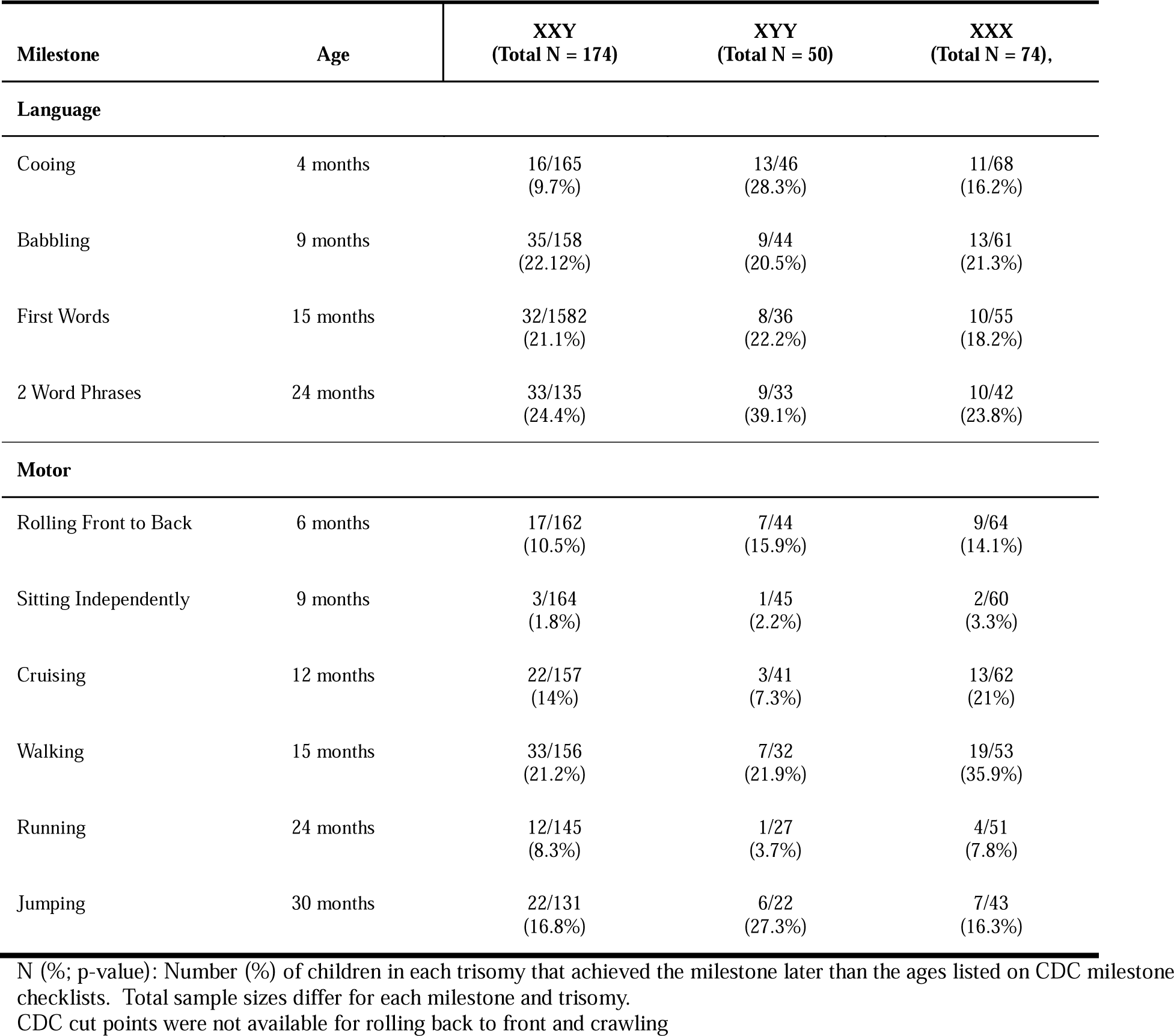
Frequencies of children with SCT delayed in milestones according to ages set by CDC Milestones checklists.

### Consideration of Early Intervention Therapies

Of the 298 children included, 187 (63.8%) had received EI therapy, started either proactively due to risk for delays or in response to developmental concerns in one or more developmental domains. There were no differences in therapy rates between the SCT conditions. Within our cohort, children with history of EI achieved milestones significantly later than children who had not (p<0.001 for all milestones). This is likely because those with identified delays were more likely to be referred for developmental therapies.

## Discussion

This study represents the first report on developmental milestone achievement in prenatally identified SCT and provides a novel milestone chart that can help parents and professionals better quantify and visualize what “increased risk for developmental delay” means in SCT conditions. These cohorts were not referred for any concerns and thus were as close to “population based” as possible. In all SCT conditions, there was a later median age of skill development across multiple gross motor and expressive language milestones than reported in normative datasets. This includes both early milestones such as cooing and rolling, and later milestones including 2-word phrases, walking, and running. Furthermore, there was more variability in the age range for milestone achievement in our sample compared with reference norms, with the range of acquisition for all milestones extending later in life for children with SCT. These findings support the need to consider infants with SCT as a group at increased risk for delays and deserving of closer developmental monitoring given that age of early motor and language milestones have been shown to predict longitudinal outcomes across all developmental domains in the general population and clinical samples.^15–26^

These results confirm prior research indicating increased risk for developmental delays in children with SCT,^4,27,28^ consistent with findings in other genetic disorders where milestone acquisition is different than population norms.^29^ However, unlike other genetic conditions such as DS and FXS,^30,31^ our results also show a significant amount of overlap with the general pediatric population. Figure 1 shows that, for many children with prenatally identified SCT, early milestones present within, or close to, the expected timeline. While this is reassuring, there are known later risks in SCT for many neurodevelopmental diagnoses including speech-language disorders, learning disabilities, ADHD, executive dysfunction, motor skill deficits, and autism spectrum disorders,^32–44^ which all benefit from earlier diagnosis and evidence-based treatments. Thus, careful attention to development trajectories is warranted as early interventions may help minimize these morbidities.

The variability of the phenotype and overlap with the general population often leads to questions of whether different developmental care pathways and extra developmental testing is needed for *all* infants with SCT. This is a valid concern as a relatively high proportion of individuals with SCT conditions have minimal neurodevelopmental differences with positive adult outcomes,^45–47^ and many go undiagnosed from their clinical presentation. Additional recommendations for developmental monitoring and evaluation may increase family stress, negatively impact parent-child relationships, and call unnecessary attention to the genetic differences in their child, as well as increase healthcare utilization and demand on a stressed early intervention system.

Prospective longitudinal research is needed to clarify if indeed there are specific early risk factors predictive of poorer outcomes that would warrant stratifying children with SCT into different low versus high-risk developmental care pathways, similar to extensive work done in the congenital heart disease and prematurity populations.^48,49^ These pathways, however, were developed using evidence from hundreds of studies, which do not currently exist in SCT. Thus, until more prospective data is available, consideration of all infants with SCT as high risk is warranted.

Table 3 responds to our interest in whether recently published milestones from the CDC^7^ are appropriate for developmental surveillance in infants with SCT. Overall, a relatively small proportion of children in our sample were delayed in milestone achievement according to CDC milestones checklists (Table 3) even though their milestone acquisition was delayed as compared to other metrics (Denver II; WHO). This suggests that relying on the CDC milestone lists for SCT will fail to identify many infants with delayed milestones and is consistent with other published concerns^50–52^ about low sensitivity of the ages presented in the CDC milestones. It is well recognized that standard developmental screening tools designed for the general population (e.g., ASQ, PEDS)^53–55^ have lower sensitivity in high-risk groups, which has led to guidelines for developmental follow-up of high-risk neonates with periodic direct assessment.^49,56–58^ Similarly, our findings of the increased risk in SCT support that periodic direct developmental assessment should be part of SCT treatment guidelines.^59^

By offering detailed information on milestone achievement, we provide a valuable tool for clinicians and families to better interpret a child’s early development within the context of their SCT condition, rather than only comparing to general population norms. Further, any significant deviations from SCT norms may alert clinicians to potential risks for comorbid health conditions or an additional genetic difference. While pediatric providers can use this tool as a reference to contextualize a child’s milestone achievement, it is not intended to delay referrals for developmental evaluations or early intervention support. Parents may appreciate the more nuanced normative data as they track their child’s milestones, noting areas where their child’s development aligns with children with similar genetic profiles as well as areas of normative differences. Prior research shows parents of children with delayed milestones may have higher levels of perceived stress^60^ or experience guilt that they have done something to cause their child’s delays.^61^ A clearly defined schedule for the timing of developmental milestones specific to each SCT, when used in conjunction with normative milestones expectations, may be more palatable in supporting early developmental care.

Results showing similarities and differences in milestone achievement by karyotype (Table 2) add to the existing literature on genetic disorders by providing more specific data regarding milestone acquisition in each trisomy condition. For most milestones, SCT groups were statistically similar. This aligns with prior research showing similar early developmental and neurocognitive profiles across the SCT conditions.^44,62–64^ However, the XXY group did achieve several milestones earlier, including cooing 1 month earlier than both other groups and crawling and cruising 1 month earlier than those with XXX. While this may be an artifact of a larger and more variable sample size in XXY, it may also reflect differential effects of the extra X chromosome in males.^65^ Ongoing research with larger sample sizes for XYY and XXX will help determine if different SCT conditions have clinically relevant differences in developmental trajectories.

These study results also have practical implications for genetic counseling and are responsive to prior research findings showing that parents receiving a prenatal SCT diagnosis desire more accurate and current data on potential neurodevelopmental outcomes specific to each SCT condition. In the context of highly variable phenotypes associated with SCT, genetic counselors strive to provide guidance to parents with a new diagnosis and clarify parental perception of risks for developmental delays.^66^ This foundation establishes how parents understand and respond to their child’s development and behavior, especially as related to the genetic diagnosis. By providing a clearer picture of developmental expectations associated with the diagnosis, genetic counselors can more specifically inform parents about what to expect in their child’s first few years of life, as well as promote awareness, empowerment, and a proactive approach to early intervention processes to facilitate early developmental care.^67^

Despite the insights gained, limitations are important to consider. First, smaller sample sizes for the XXX and XYY karyotypes limit the generalizability compared with the XXY sample.

Additionally, there are known limitations in normative data for milestone acquisition, including unclear and inconsistent definitions of milestones,^68^ ambiguity around what constitutes achievement of the milestone (partial vs. complete),^69^ and differences in the raters used to determine milestone achievement for normative datasets (parents vs. clinicians).^68,70,71^ Further, normative datasets rarely account for potential sex differences,^72,73^ racial and sociocultural differences,^50,74,75^ and known variability related to social determinants of health^69^ (SDoH), which further adds a degree of uncertainty to our findings. Our sample was disproportionately white, non-Hispanic with high socioeconomic status, and future studies should aim to include more representative samples. Parental recall bias is another commonly recognized challenge when evaluating parent-reported milestones,^76,77^ however minimized in this study with frequent visits at 2, 6, 12, 18, 24, and 36 months of age with pediatric physicians interviewing and verifying milestone achievements. Importantly, while there are many benefits to an ongoing natural history study, our study design is limited in that at the time of publication, not all participants in the sample had yet achieved all 12 milestones measured and therefore sample sizes were different for each SCT condition at each milestone. Additionally, while we explored the effect of early intervention in our analysis, the act of participating in a natural history study itself may impact developmental course. Families in the study have self-selected to participate in regularly scheduled developmental evaluations with expert SCT clinicians and to monitor developmental milestones in between visits. While a prenatally identified sample of nearly 300 infants with SCT provides a less biased dataset than prior studies, it may still not fully represent the broad spectrum of outcomes in SCT. Future results based on direct assessments through the eXtraordinarY Babies study can address these limitations and further refine our understanding of developmental trajectories and risk groups in this population.

In conclusion, developmental milestone achievement in SCT conditions is delayed compared to the general population, however only in a subset of infants with SCT. As increasing numbers of infants with prenatal SCT diagnoses present at pediatric practices, we provide an evidence-based schedule of milestone achievement in SCT as a tool for families, pediatricians, genetic counselors, and early intervention teams. Utilization of such a tool can support shared clinical decision-making between parents and providers, promoting timely referrals and identifying patterns inconsistent with SCT. However, given the paucity of prospective research identifying specific risk factors for later negative outcomes, recommended care for SCT conditions should follow practices of other high-risk conditions - with more responsive attention to developmental concerns, recognition that standard surveillance and screening tools have lower sensitivities in high-risk populations, and referrals for periodic direct developmental assessments. While more rigorous research will help identify evidence for timing of direct assessments and highest-risk groups, general publications support assessments at 6-12 months, 18-24 months, and 36 months of age.^78–80^

## Data Availability

All data produced in the present study are available upon reasonable request to the authors

## Author contributions

- Conceptualization - Talia Thompson, Nicole Tartaglia

- Statistical analysis - Samantha Bothwell

- Data collection - Jennifer Janusz, Rebecca Wilson, Susan Howell, Nicole Tartaglia, Shanlee Davis, Judy Ross, Karen Kowal, Chijioke Ikomi, Talia Thompson

- Critical reviewing of the manuscript - Jennifer Janusz, Rebecca Wilson, Susan Howell, Shanlee Davis, Karli Swenson, Sydney Martin, Judith Ross, Maria Despradel

- Data cleaning - Maria Despradel

## Abbreviations

SCT: Sex Chromosome Trisomy
CDC: Centers for Disease Control
AAP: American Academy of Pediatrics
DS: Down syndrome
FXS: Fragile X syndrome
CNS: Central Nervous System
WHO: World Health Organization
PRP: Primitive Reflex Profile
SDoH: Social Determinants of Health
EI: Early Intervention

